# MRI Findings of Myocardial Fibrosis among Patients Living With HIV: A Systematic Review and Meta-Analysis

**DOI:** 10.64898/2026.09.23.26363040

**Authors:** Saeed Shoar, Reza Khademi, Maria Patarroyo – Aponte, Ankit Nahata, Maya Isabella Hoffman, Zachary Pinchover, William C. Harding, Gabriel Patarroyo – Aponte

## Abstract

HIV infection significantly elevated the odds of cardiovascular diseases, including myocardial fibrosis. We aimed to determine the difference of late gadolinium enhancement (LGE), extracellular volume (ECV), T1, and T2 using cardiac magnetic resonance imaging (CMR) in patients living with HIV (PLWH) and healthy controls. The databases Scopus, Web of Science, Embase, and PubMed were searched systematically using relevant keywords. STATA version 17 was used for meta-analysis using random effect and fixed effect models for heterogeneous and non-heterogeneous analyses. An overall number of 13 studies were included in the current analysis involving 1425 PLWH and 766 healthy controls. PLWH showed significantly higher LGE (OR: 5.47, 95% CI: 1.90-15.64, p<0.001) and ECV (mean difference: 1.46, 95% CI: 0.21-2.71, p=0.02) compared to controls. However, differences in Left Ventricular Ejection Fraction (LVEF) were not significant (mean difference: −1.41, 95% CI: −2.81- −0.01, p=0.05). There were no meaningful disparities in pro B-type natriuretic peptide (BNP) values (mean difference: 8.67, 95% CI: 0.04-17.29, p=0.05) or T2 values (mean difference: 0.91 ms, 95% CI: −1.04-2.86, p=0.36), but T1 values were significantly higher in PLWH (mean difference: 13.78 ms, 95% CI: 9.16-18.40, p<0.001). In meta regression, LGE was correlated with nadir CD4 (z=-3.41, p-value<0.001). In addition, ECV was correlated with white race (Z=-3.97, p-value<0.001), Hepatitis C (Z=-2.08, p-value=0.037), and smoking (Z=2.05, p-value=0.041). HIV is related to a significantly elevated rate of myocardial fibrosis, and factors such as lower nadir CD4, white race, hepatitis C, and smoking were correlated with hepatitis C.

## 1. Introduction

Human immunodeficiency virus (HIV) prognosis has substantially improved due to antiretroviral therapy (ART), leading to a life expectancy that is almost equivalent to that of individuals without HIV. However, there is a higher occurrence of severe non-acquired immune deficiency syndrome (AIDS)-related illnesses, such as cardiovascular diseases ^1, 2^. By 2030, projections indicate that over 70% of people living with HIV (PLWH) will be aged 50 years or older, and 78% of PLWH are expected to have a diagnosis of cardiovascular disease ^3^. PLWH have an elevated likelihood of having cardiovascular diseases, including heart failure (HF) and sudden cardiac death (SCD), in comparison to the overall population ^4^. The exact cause of HF in PLWH is not completely understood ^5^. Research indicates that PLWH currently show a frequency of HF with preserved ejection fraction ranging from 15% to 30%, and an occurrence of subclinical cardiomyopathy with echocardiographic indications of diastolic dysfunction approaching 50% ^6^. While it is more prevalent after coronary diseases in PLWH ^7^, the higher occurrence of HF in PLWH cannot be simply attributed to the increased likelihood of coronary diseases ^2^. Several different mechanisms, such as myocarditis and myocardial fibrosis, have been suggested as potential causes ^8, 9^. Cardiac magnetic resonance imaging (CMR) is providing growing evidence that myocardial inflammation, fibrosis, and steatosis are involved in the process ^10, 11^. Myocardial fibrosis is a multifaceted course that can be initiated by injury, stress, and inflammation, leading to the buildup of extracellular matrix in the myocardium ^12^. It was previously shown that the rate of diffuse intramyocardial fibrosis, as measured by CMR, is higher in PLWH without known cardiovascular disease, and CMR could be a valuable marker for detecting subclinical myocardial injury in this population ^9^. The late gadolinium enhancement (LGE) pattern and extracellular volume fraction (ECV), detected by CMR, indicate regional fibrosis, which might signify permanent damage to the heart muscle and lead to impaired cardiac function and potentially fatal cardiac events ^13–15^. We aim to assess the difference in prevalence of myocardial fibrosis in PLWH and healthy subjects, as well as the factors that are correlated with the occurrence of myocardial fibrosis.

## 2. Methods

### 2.1 Data search

We followed the Preferred Reporting Items for systematic reviews and meta-analyses (PRISMA) standards when designing, conducting, and reporting our systematic review. The protocol of this study was published before ^16^. PubMed, Embase, Web of Science (WOS), and Scopus were searched for relevant articles until June 9^th^, 2025. The strategy used for the search was as follows: (“HIV” OR “human immunodeficiency virus”) AND (“Cardiac fibrosis” OR “myocardial fibrosis” OR “endomyocardial fibrosis” OR “late gadolinium enhancement” OR “LGE” OR “delayed enhancement” OR “endocardial fibrosis”) and **Supplementary Table 1** contains the complete search strategies used for each database. This systematic review and meta-analysis synthesized data from previously published studies. All included studies were assumed to comply with ethical guidelines as stated in their respective publications. This research was carried out following the ethical guidelines of the Declaration of Helsinki.

### 2.2 Inclusion and Exclusion Criteria

The inclusion criteria of this study were all studies that use CMR and compare the rate of myocardial fibrosis in adult PLWH and healthy controls. Single-arm studies, studies on autopsy, studies on individuals younger than 18 years old, reviews, case reports, conference abstracts, letters, and Articles written in languages other than English were excluded.

### 2.3 Outcome Measures

The key objective of this research was to compare the LGE and myocardial ECV as markers of myocardial fibrosis between PLWH and healthy controls. In addition, the secondary outcomes were to compare T1, T2, pro-brain natriuretic peptide (BNP), and left ventricular ejection fraction (LVEF) between these two groups. Furthermore, meta-regression was conducted to find correlated factors with LGE and ECV differences between PLWH and healthy controls.

### 2.4 Study Selection

After duplicates were removed, two independent authors screened articles by title and abstract. After screening the title and abstract, the entire paper was reviewed separately to determine if it met all the eligibility criteria. Discrepancies were settled by a third author after reaching a consensus. Furthermore, a manual search was conducted by examining the references of selected articles.

### 2.5 Quality Assessment

Two separate investigators evaluated the credibility of the included articles during data extraction by the Newcastle-Ottawa Scale ^17^. Any discrepancies were addressed by consulting a third reviewer and reaching a consensus. Studies were divided into high (7-9 stars), moderate (5-6 stars), and low quality (less than 5 stars) based on the Newcastle-Ottawa score.

### 2.6 Data Extraction

Two independent reviewers acquired the subsequent variables from the selected papers, including author, Study design, inclusion and exclusion criteria, sample size, age, gender, country, year of publication, race, comorbidities, laboratory investigations, duration since HIV diagnosis, using ART, time since ART use, CD4+ T-cell count, and main and secondary outcomes of our study. In terms of any discrepancies in extracted data, full texts were reviewed again by a third reviewer. If there were any uncertainties regarding the variables, the corresponding author of the study was contacted.

### 2.7 Data synthesis

The data were obtained in the form of the mean and standard deviation. The provided information includes the median and interquartile range, as well as the mean and range, for some variables. The formula developed by Hozo et al., Luo et al., and Wan et al. was used to transform these variables into their respective mean and standard deviations ^18–20^. The analysis was conducted by Stata/SE, version 17, developed by StataCorp LLC. The I^2^ statistic was utilized to compute heterogeneity, and studies with an I^2^ value exceeding 50% were deemed to exhibit significant heterogeneity. Variables with meaningful heterogeneity were assessed by a random effects model, whereas variables with minimal heterogeneity were interpreted using a fixed effects model. Random effect meta-regression was used to find any correlation between main outcomes differences and other variables.

### 2.8 Publication bias

To assess potential publication bias of our meta-analyses, we conducted a funnel plot and trim-and-fill analysis, which was employed to identify any potentially missing studies in cases of funnel plot asymmetry.

### 2.9 Patient and Public Involvement

Patients and members of the public were not involved in the design, conduct, reporting, or dissemination plans of this systematic review and meta-analysis.

## 3. Results

### 3.1 Search results

The initial search of databases yielded a total of 570 articles. After excluding 261 duplicated articles, 309 titles and abstracts of articles were screened. After eliminating 235 unrelated articles, 74 articles underwent full-text screening. Finally, a total of 13 studies met our inclusion criteria and were included, and no studies were retrieved via manual search, as **Figure 1** demonstrates the selection process in detail. **Table 1** and **2** shows the study and population features of the selected research.

**Figure 1:**
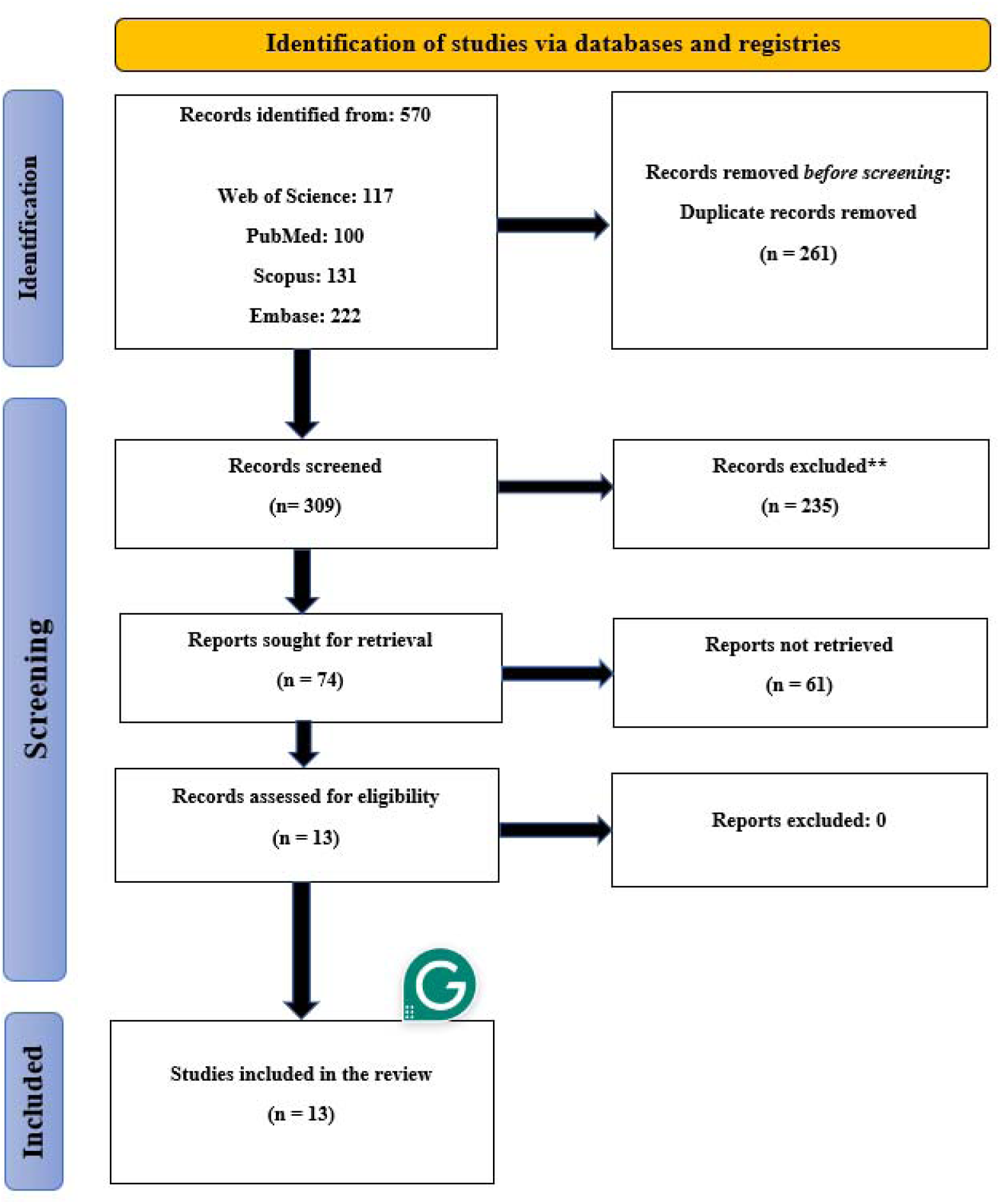
The PRISMA flowchart of included studies.

**Table 1:**
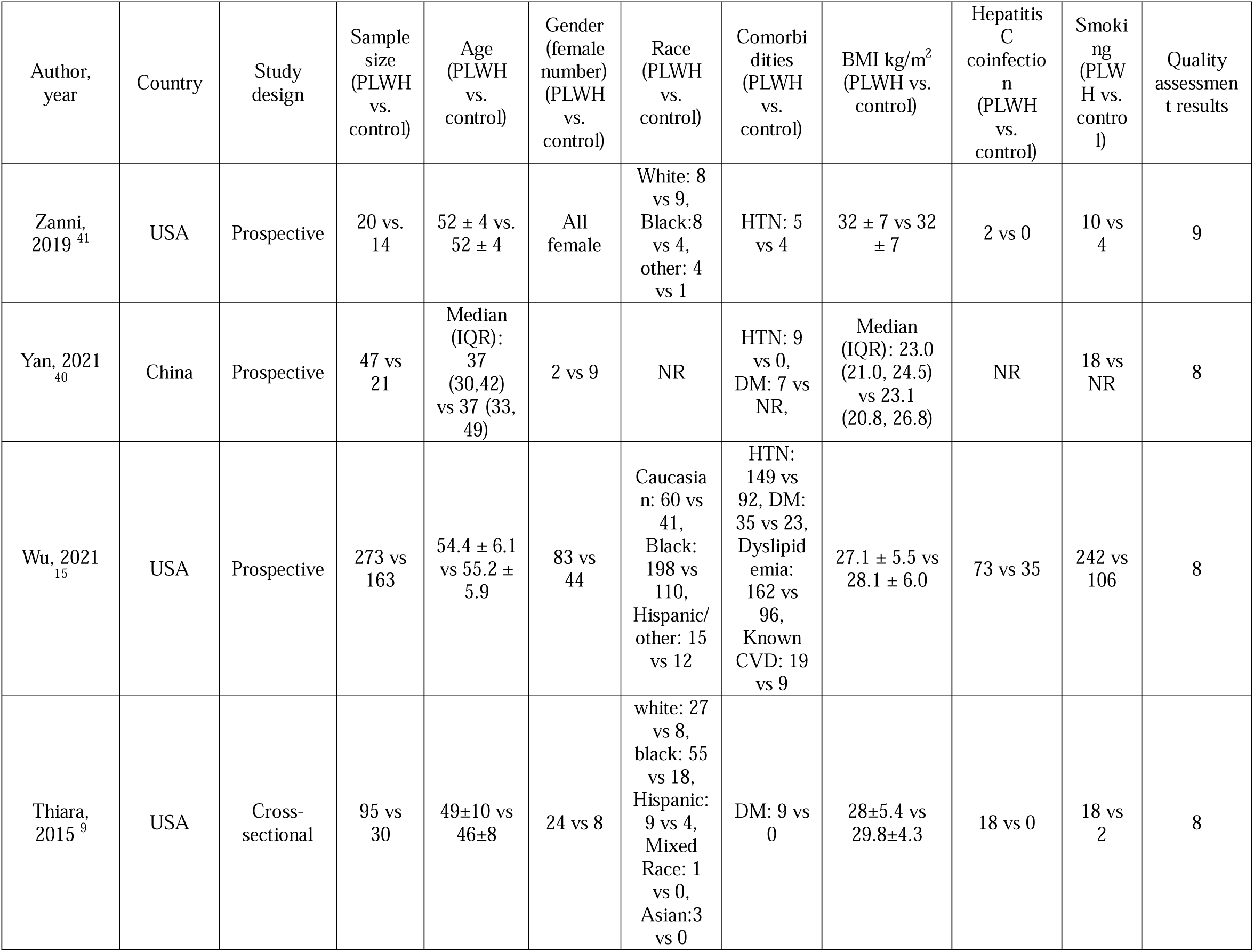
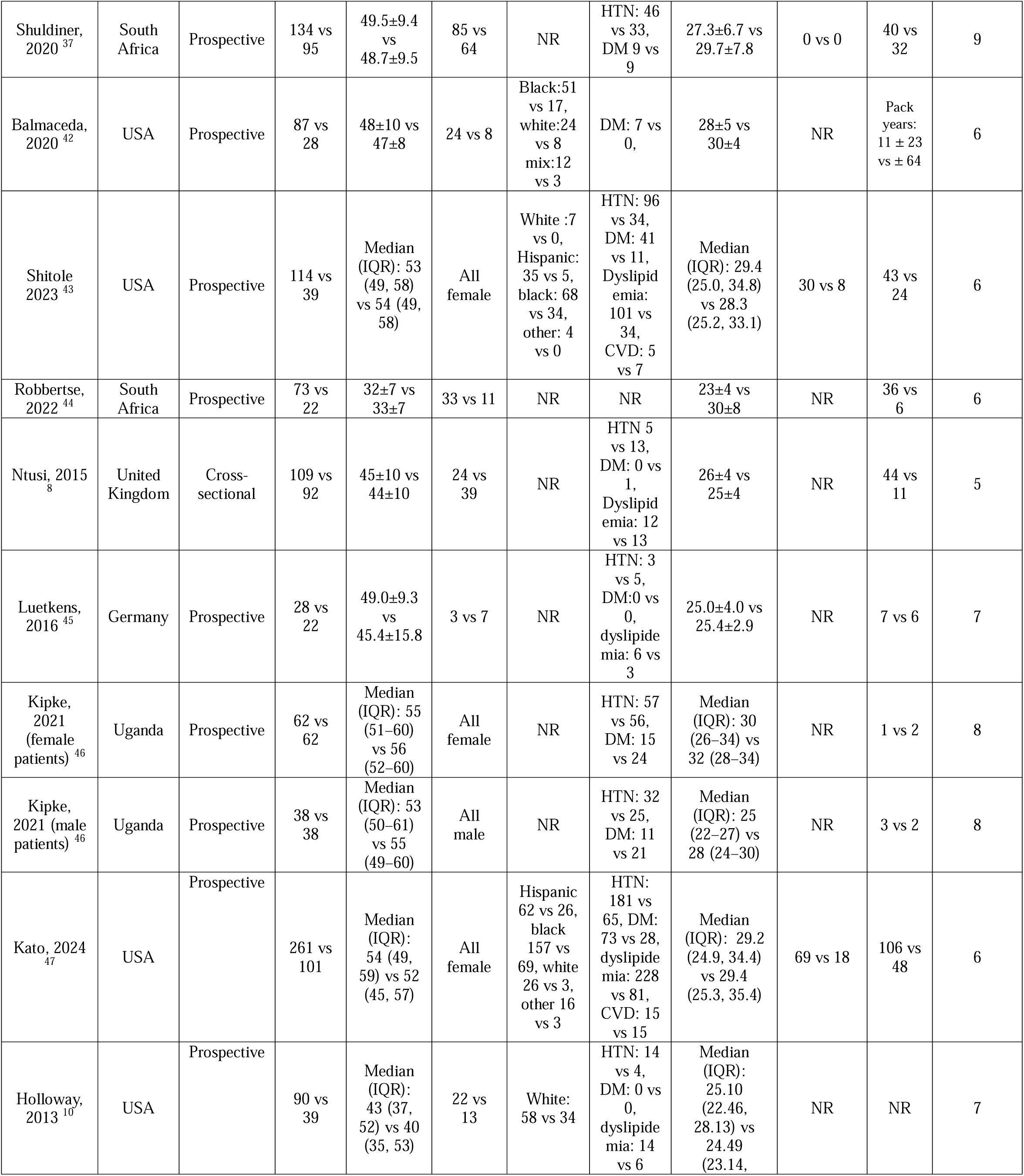
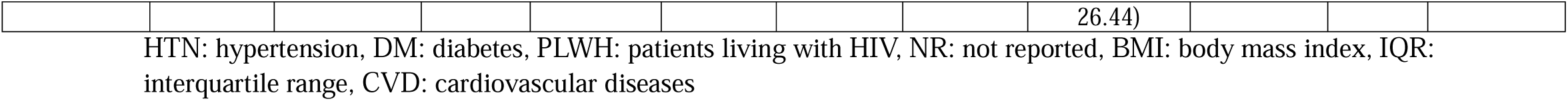
The baseline characteristics of included studies.

**Table 2:** Population characteristics of included studies.

| Author, year | Time since HIV diagnosis (year) | Total duration of ART (years) | ART use | CD4+ T-cell count, cells/μL | Nadir CD4+ T-cell count, cells/μL |
| --- | --- | --- | --- | --- | --- |
| Zanni, 2019 | 19 ± 8 | Median (IQR): 18 (8–22) | All patients | median (IQR): 773 (526–1202) | median (IQR): 138 (18–270) |
| Yan, 2021 | Median (IQR): 3.4 (1.4, 7.3) for HIV and 6.0 (5.0, 11.5) for Aids | Median (IQR): 2.5 (1.5, 5.1) for HIV and 1.5 (0.3, 6.3) for AIDS | 37 | 717.2±209.8 for HIV and 188.2±92.0 for AIDS | NR |
| Wu, 2021 | NR | 10.8± 6.1 years | 240 | Median (IQR): 607 (393–826) | Median (IQR): 270 (141–400) |
| Thiara, 2015 | 14±8 | 9±6 | 88 | 615±276 | 235±182 |
| Shuldiner, 2020 | 9.5±5.8 | 7.0±4.3 | All | 534±270 | 271±213 |
| Balmaceda, 2020 | NR | NR | NR | 607±277 | 0.81±0.56 |
| Shitole 2023 | NR | NR | 112 | Median (IQR): 665 (489, 854) | Median (IQR): 172 (84, 267) |
| Robbertse, 2022 | 8 (5 to 26) days | NR | All | 289 (170 to 410) | NR |
| Ntusi, 2015 | NR | NR | 90 | NR | NR |
| Luetkens, 2016 | 9.7±6.9 | NR | All | 475.1±307.9 | 129.0±126.7 |
| Kipke, 2021 (female patients) | Median (IQR): 12 (10–12) | Median (IQR): 11 (9–12) | 62 | Median (IQR): 475.1±307.9 | Median (IQR): 129.0±126.7 |
| Kipke, 2021 (male patients) | Median (IQR): 12 (10–13) | Median (IQR): 11 (9–13) | 38 | Median (IQR): 593 (504–718) | Median (IQR): 141 (75–199) |
| Kato, 2024 | NR | NR | 243 | Median (IQR): 700 (500, 925) | NR |
| Holloway, 2013 | NR | NR | All | 546±179 | NR |

NR: not reported, AIDS: acquired immunodeficiency syndrome, IQR: interquartile range, ART: antiretroviral therapy

### 3.2 Study characteristics

The study design of all the included studies was observational. A total of 1425 PLWH and 766 healthy controls were included in the current study. The geographical distribution of published articles was as follows: USA (7 articles), South Africa (2 articles), Uganda (1 article), United Kingdom (1 article), Germany (1 article), and China (1 article). The PLWH group included 757 (53.12%) female patients, while the control group had 419 (54.69%) females. The weighted mean age was similar between the two groups (PLWH: 49.73 years, control: 49.72 years). Among PLWH, 95.56% were on ART. On average, PLWH had been diagnosed with HIV for 9.9 years and had been on ART for 9.97 years. In PLWH, the mean CD4 count was 594 cells/μL, with a nadir CD4 count of 224.19 cells/μL.

### 3.3 Late gadolinium enhancement (LGE)

A total of 8 studies reported the rate of LGE in PLWH and healthy controls. The results of the random effect model pooled analysis showed that the rate of LGE was notably higher in PLWH in comparison to healthy controls (OR: 5.47, 95% CI: 1.90-15.64, p<0.001) (**Figure 2**). To assess the robustness of our meta-analysis results, we conducted a leave-one-out sensitivity analysis by a random-effects model. The results of this analysis are depicted in **Supplementary Figure 1**. The results suggest that no single study disproportionately influences the overall effect size. Furthermore, the p-values for each analysis (all <0.01) confirm that the treatment effect remains statistically significant even when any one study is excluded.

**Figure 2:**
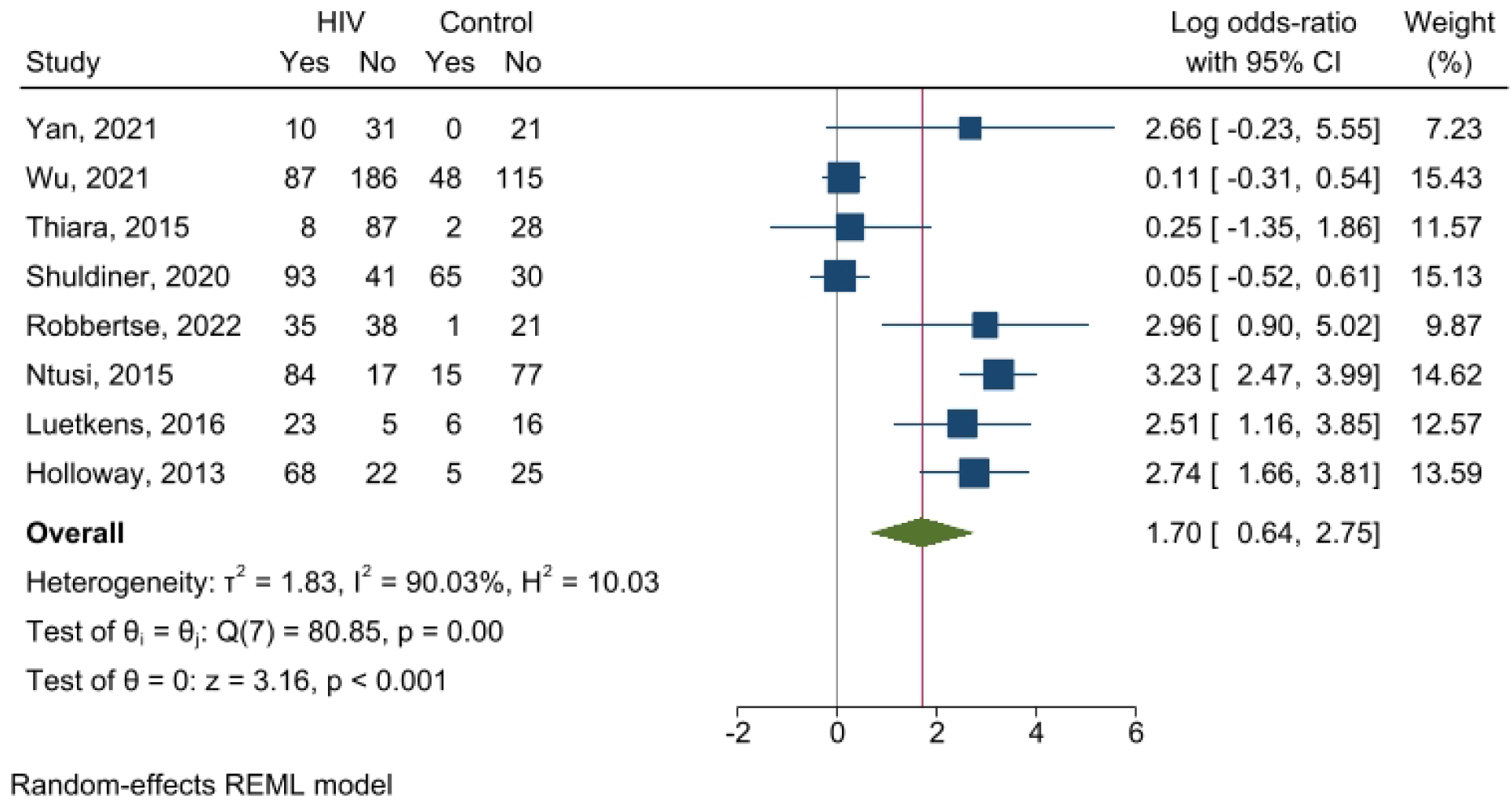
The results of pooled analysis of LGE between PLWH and healthy controls in a pooled analysis of 8 studies.

### 3.4 Myocardial extracellular volume fraction (ECV)

Myocardial ECV was reported by 8 studies. The random effect pooled analysis showed that PLWH had significantly higher ECV with a mean difference of 1.46 in comparison to healthy controls (95% CI: 0.21-2.71, p=0.02) (**Figure 3**). The result of the leave-one-out sensitivity analysis for ECV is shown in **Supplementary Figure 2**. However, the exclusion of Markella V. Zanni (2019), Yan (2021), Balmaceda (2020), and Robbertse (2022) results in p-values that were not significant. This suggests that while the overall effect remains consistent, the statistical significance of the results is somewhat sensitive to the exclusion of these studies.

**Figure 3:**
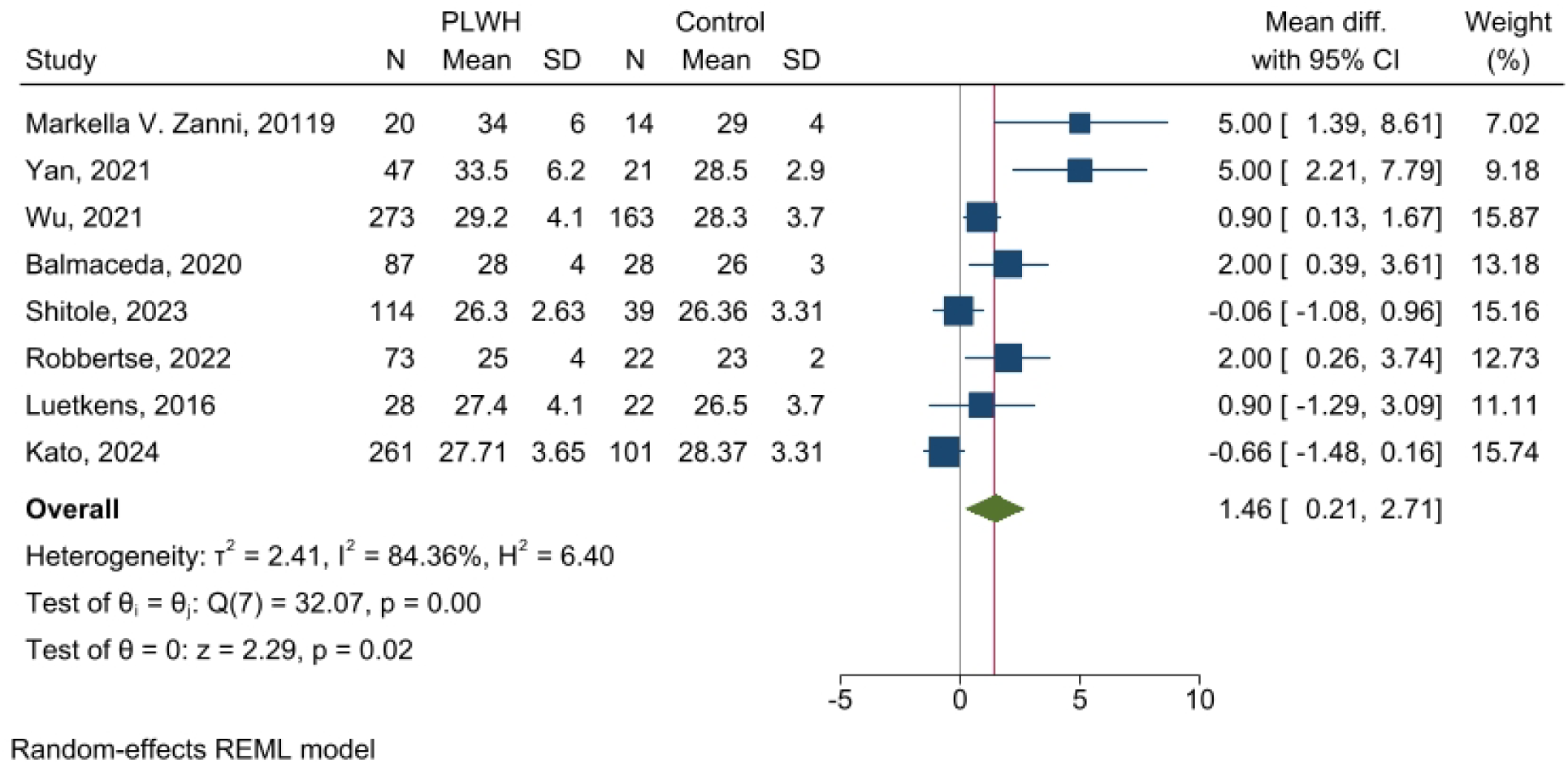
The results of pooled analysis of ECV between PLWH and healthy controls in a pooled analysis of 8 studies.

### 3.5 T1 and T2

The fixed effect pooled analysis of 7 studies showed that patients with PLWH had significantly higher T1 compared to healthy controls (mean difference: 13.78 ms, 95% CI: 9.16-18.40, p<0.001) (Supplementary Figure 3). Furthermore, the random effect pooled analysis of the two research showed that T2 was not notably different among PLWH and healthy controls (mean difference: 0.91 ms, 95% CI: −1.04-2.86, p=0.36) (Supplementary Figure 4).

### 3.6 LVEF and Pro-BNP

As shown in Supplementary Figures 5 and 6, LVEF and Pro-BNP were not markedly different among PLWH and healthy controls (mean difference: −1.41, 95% CI: −2.81- −0.01, p=0.05 and mean difference: 8.67, 95% CI: 0.04-17.29, p=0.05, respectively).

### 3.7 Meta regression

In meta regression, LGE was significantly correlated with nadir CD4 (z=-3.41, p-value<0.001). In addition, ECV was significantly correlated with percentage of white race (Z=-3.97, p-value<0.001), Hepatitis C (Z=-2.08, p-value=0.037), and smoking (Z=2.05, p-value=0.041). LGE and ECV were not correlated with the percentage of patients who were on ART.

### 3.8 Publication bias

The funnel plots of ECV and LGE are shown in Supplementary Figures 7 and 8, The funnel plots show asymmetric distribution of studies in which a larger number of studies fall to the right of the overall estimated effect size compared to the left. These results suggest a potential for publication bias, where research demonstrating positive outcomes tends to have a higher publication likelihood and is included in the analysis. However, trim-and-fill analysis did not show changes in outcomes.

### 3.9 Quality assessment

The findings of the quality evaluation of the included research are shown in **Table 1** and supplementary Table 2. All the included studies had moderate and high quality based on the Newcastle-Ottawa score.

## 4. Discussion

In the current study, we conducted a systematic review and meta-analysis to explore the correlation of myocardial fibrosis and HIV. A total of 13 studies with 1425 PLWH and 766 healthy controls were included. Our results showed that PLWH had a notably elevated rate of LGE, ECV, and T1 compared to healthy controls.

Myocardial fibrosis happens on a spectrum ranging from mild to severe, characterized by the presence of excessive collagen in the cardiac interstitium ^21^. Consequently, Myocardial native T1 and ECV increase, but post-contrast T1 declines ^22^. ECV is highly effective for assessing interstitial expansion associated with fibrosis using extracellular gadolinium-based contrast agents. ECV is capable of identifying subtle interstitial expansion that is not easily detected by LGE. It has been demonstrated that ECV can predict clinical outcomes with a same level of significance as LVEF when assessing prognosis ^23, 24^. ECV quantifies the interstitial concentration of gadolinium relative to plasma levels. Histological validation data consistently demonstrate superior agreement with ECV compared to other T1 metrics, as indicated by higher R2 values, reflecting the proportion of variation explained by another variable ^25–27^. Our results showed that PLWH had 1.46% and 13.78 ms higher ECV and T1 relaxation time compared to healthy controls, respectively. Two recent extensive studies on patients without HIV who were referred for CMR examinations discovered a direct correlation between higher ECV levels and adverse outcomes ^28, 29^. Each increase of 4-5% (1 standard deviation) in ECV was linked to a hazard ratio (HR) of 1.28-1.41 for the likelihood of being hospitalized or a mortality due to HF ^28, 29^.

Furthermore, in the context of subclinical myocarditis, the presence of LGE may indicate permanent damage to the heart muscle, consistent with observations made in people without HIV ^13^. The existence of LGE is related with a poorer prognosis compared to its absence ^30^. Our results showed that PLWH had 5.47 times higher rate of LGE in comparison to healthy controls. In addition, in line with our results, Tseng et al compared HIV and non-HIV patients who had sudden cardiac arrest and underwent autopsy. They showed higher histologic levels of interstitial myocardial fibrosis in PLWH relative to non-HIV patients ^31^.

Furthermore, our findings contribute to the expanding body of evidence suggesting that, alongside myocardial edema, PLWH may also exhibit features of replacement-type myocardial fibrosis. HIV infection treated with combination ART is linked with an elevated occurrence of myocardial fibrosis, and impaired left ventricular function during both systole and diastole ^32–34^. However, effective ART has shifted the main manifestation of HF in PLWH from left ventricular systolic dysfunction to left ventricular diastolic dysfunction ^35^. In a previous systematic review and meta-analysis done by Zhang et al, it was shown that over a median follow-up period of 31.2 months, higher levels of cardiac fibrosis were significantly associated with an elevated odds of major adverse cardiovascular events (MACE) (HR = 1.34, 95% CI = 1.14–1.57) and all-cause mortality (HR = 1.74, 95% CI = 1.27–2.39) ^36^. Therefore, these findings underscore the importance of monitoring PLWH to detect high risk patients and mitigate the odds of adverse cardiovascular consequences.

Furthermore, the association of using ART and cardiac fibrosis in PLWH is not clearly understood yet. Although one previous study showed no significant correlation between ART use and myocardial fibrosis ^37^, another study indicated that ART-naive individuals had elevated levels of myocardial fibrosis compared to those with established HIV treatment regimens ^38^. In addition, a previous systematic review done by Hudson et al. showed a notable and gradual correlation between raised CD4 cell counts and decreased rates of myocardial fibrosis ^39^. Although our meta-regression analysis did not show a correlation between the percentage of ART use and myocardial fibrosis indices, it was shown that higher LGE presence was correlated with lower nadir CD4 count. In line with our results, Yan et al reported that ECV values and the presence of LGE increased with the progression of HIV, indicating a connection between the clinical severity of HIV and cardiac fibrosis. Additionally, AIDS was identified as a risk factor for myocardial fibrosis, reinforcing the recommendation that all HIV patients should start ART to manage disease progression ^40^.

Our study outcomes should be considered alongside some limitations. First, the heterogeneity of some meta-analyses was high, which can be due to differences in baseline characteristics and risk factors among included patients, different disease severity, time from HIV diagnosis, and medications. However, meta-regression analysis was conducted to find the source of heterogeneity. Secondly, all the included research was observational, which increases the susceptibility to selection bias. Furthermore, the results of meta-regression analysis showed a significant correlation between ECV and percentage of white race, hepatitis C, and smoking, which underscore the importance of further investigation on racial differences, co-infection of hepatitis C and HIV, and other risk factors like smoking on the presence of myocardial fibrosis and its clinical presentations of PLWH.

## 5. Conclusion

In conclusion, patients living with HIV have significantly higher ECV, T1, and LGE in comparison to healthy controls. These results show an elevated presence of myocardial fibrosis in PLWH. Also, it was shown that higher LGE presence was correlated with lower nadir CD4 count, which indicates a connection between the clinical severity of HIV and cardiac fibrosis and reinforces the recommendation that all HIV patients should start ART to manage disease progression.

## Abbreviations

LGE: late gadolinium enhancement
ECV: extracellular volume
CMR: cardiac magnetic resonance imaging
PLWH: patients living with HIV
LVEF: left ventricular ejection fraction
BNP: B-type natriuretic peptide
HIV: human immunodeficiency virus
ART: antiretroviral therapy
AIDS: acquired immune deficiency syndrome
HF: heart failure
SCD: sudden cardiac death
PRISMA: Preferred Reporting Items for systematic reviews and meta-analyses
WOS: Web of Science
MACE: major adverse cardiovascular events
HR: hazard ratio
CI: confidence interval

## Declarations

### Ethics approval and consent to participate

Not applicable.

### Consent for publication

Not applicable.

### Availability of data and materials

The data supporting this article are available from the corresponding author upon reasonable and justified request.

## Authors’ contributions

Saeed Shoar: Conceptualization, study design, oversight of the systematic review process, and manuscript preparation.

Reza Khademi and Maria Patarroyo-Aponte: Performed literature search across databases and conducted study selection.

Reza Khademi, Ankit Nahata, and Maya Isabella Hoffman: Conducted data extraction.

Ankit Nahata and Maya Isabella Hoffman: Performed statistical analyses, meta-analysis, and meta-regression.

Zachary Pinchover and William C. Harding: Performed quality assessment of included studies.

Gabriel Patarroyo-Aponte: Drafted sections of the manuscript and performed critical revision for important intellectual content.

All authors contributed to the interpretation of results, read, and approved the final manuscript.

## Data Availability

All data produced in the present work are contained in the manuscript

## Acknowledgements

None.

## Supplementary Tables

**Supplementary Table 1:**
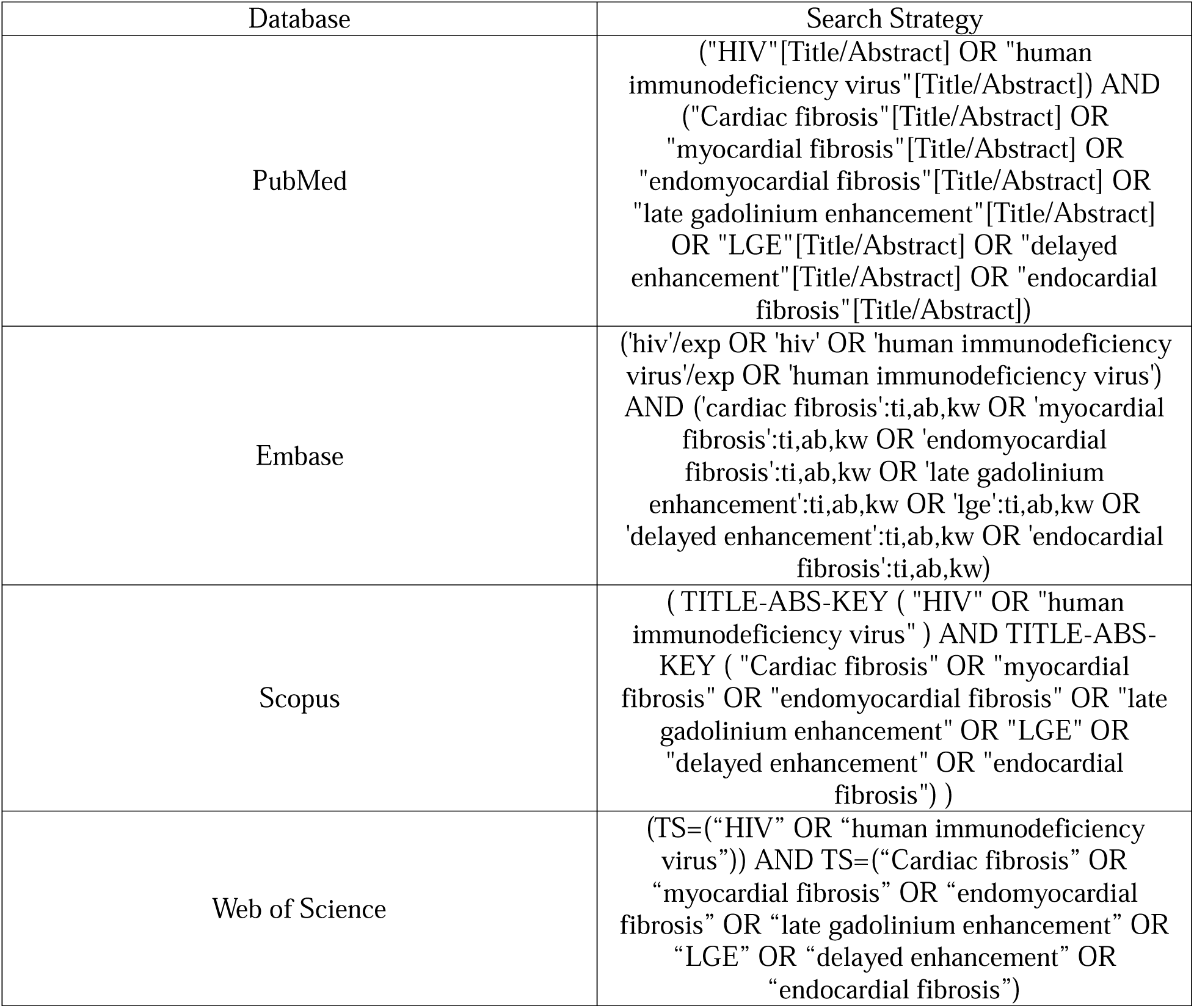
Search strategy of PubMed, Scopus, Embase, and Web of Science.

| Database | Search Strategy |
| --- | --- |
| PubMed | ("HIV"[Title/Abstract] OR "human immunodeficiency virus"[Title/Abstract]) AND ("Cardiac fibrosis"[Title/Abstract] OR "myocardial fibrosis"[Title/Abstract] OR "endomyocardial fibrosis"[Title/Abstract] OR "late gadolinium enhancement"[Title/Abstract] OR "LGE"[Title/Abstract] OR "delayed enhancement"[Title/Abstract] OR "endocardial fibrosis"[Title/Abstract]) |
| Embase | ('hiv'/exp OR 'hiv' OR 'human immunodeficiency virus'/exp OR 'human immunodeficiency virus') AND ('cardiac fibrosis':ti,ab,kw OR 'myocardial fibrosis':ti,ab,kw OR 'endomyocardial fibrosis':ti,ab,kw OR 'late gadolinium enhancement':ti,ab,kw OR 'lge':ti,ab,kw OR 'delayed enhancement':ti,ab,kw OR 'endocardial fibrosis':ti,ab,kw) |
| Scopus | ( TITLE-ABS-KEY ( "HIV" OR "human immunodeficiency virus" ) AND TITLE-ABS-KEY ( "Cardiac fibrosis" OR "myocardial fibrosis" OR "endomyocardial fibrosis" OR "late gadolinium enhancement" OR "LGE" OR "delayed enhancement" OR "endocardial fibrosis" ) ) |
| Web of Science | (TS=(“HIV” OR “human immunodeficiency virus”)) AND TS=(“Cardiac fibrosis” OR “myocardial fibrosis” OR “endomyocardial fibrosis” OR “late gadolinium enhancement” OR “LGE” OR “delayed enhancement” OR “endocardial fibrosis”) |

**Supplementary Table 2:**
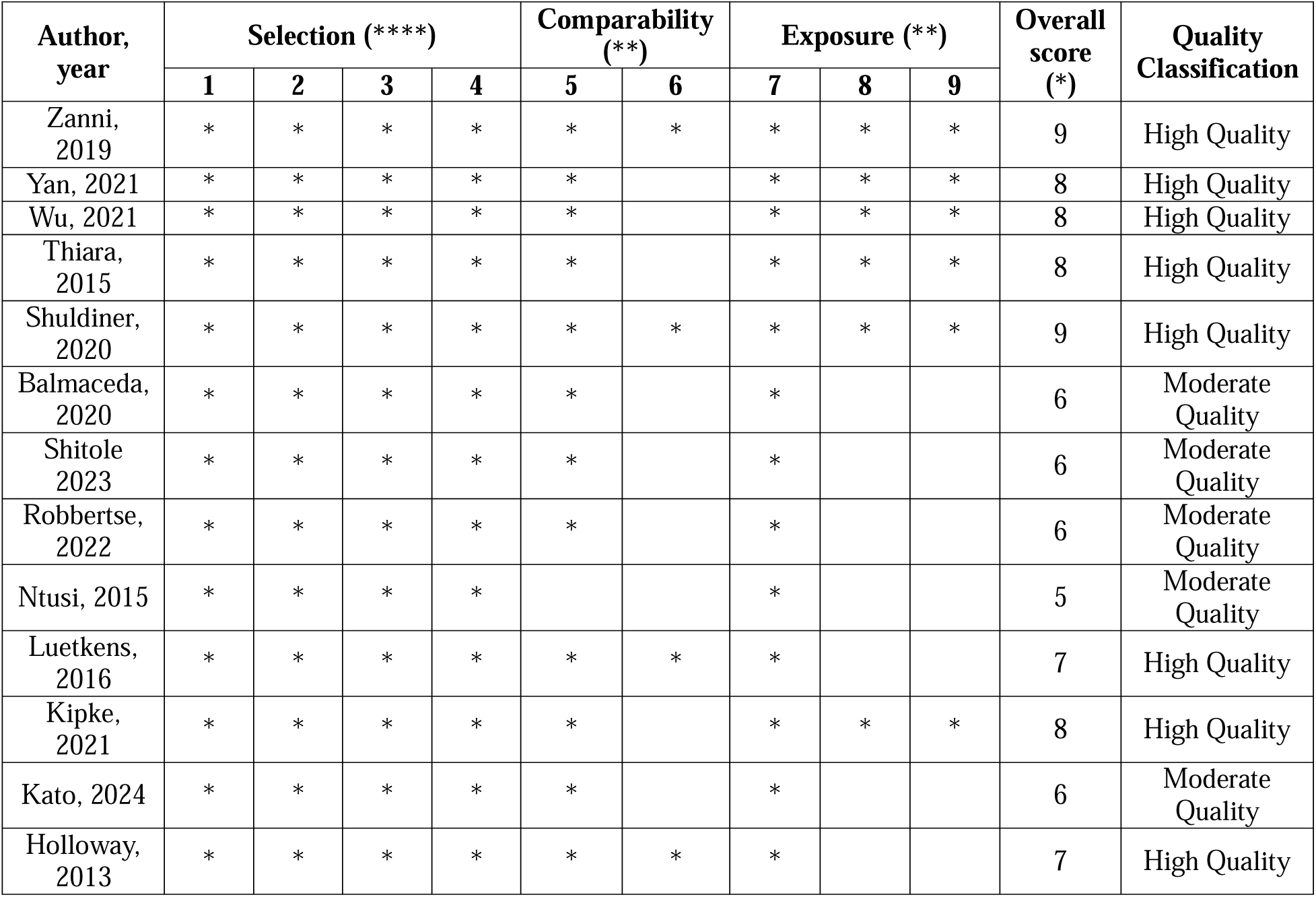
The results of the quality assessment of included studies.

| Author,<br>year | Selection (****) |  |  |  | Comparability<br>(**) |  | Exposure (**) |  |  | Overall<br>score<br>(*) | Quality<br>Classification |
| --- | --- | --- | --- | --- | --- | --- | --- | --- | --- | --- | --- |
|  | 1 | 2 | 3 | 4 | 5 | 6 | 7 | 8 | 9 |  |  |
| Zanni,<br>2019 | * | * | * | * | * | * | * | * | * | 9 | High Quality |
| Yan, 2021 | * | * | * | * | * |  | * | * | * | 8 | High Quality |
| Wu, 2021 | * | * | * | * | * |  | * | * | * | 8 | High Quality |
| Thiara,<br>2015 | * | * | * | * | * |  | * | * | * | 8 | High Quality |
| Shuldiner,<br>2020 | * | * | * | * | * | * | * | * | * | 9 | High Quality |
| Balmaceda,<br>2020 | * | * | * | * | * |  | * |  |  | 6 | Moderate<br>Quality |
| Shitole<br>2023 | * | * | * | * | * |  | * |  |  | 6 | Moderate<br>Quality |
| Robbertse,<br>2022 | * | * | * | * | * |  | * |  |  | 6 | Moderate<br>Quality |
| Ntusi, 2015 | * | * | * | * |  |  | * |  |  | 5 | Moderate<br>Quality |
| Luetkens,<br>2016 | * | * | * | * | * | * | * |  |  | 7 | High Quality |
| Kipke,<br>2021 | * | * | * | * | * |  | * | * | * | 8 | High Quality |
| Kato, 2024 | * | * | * | * | * |  | * |  |  | 6 | Moderate<br>Quality |
| Holloway,<br>2013 | * | * | * | * | * | * | * |  |  | 7 | High Quality |

## Supplementary Figures

**Supplementary Figure 1:**
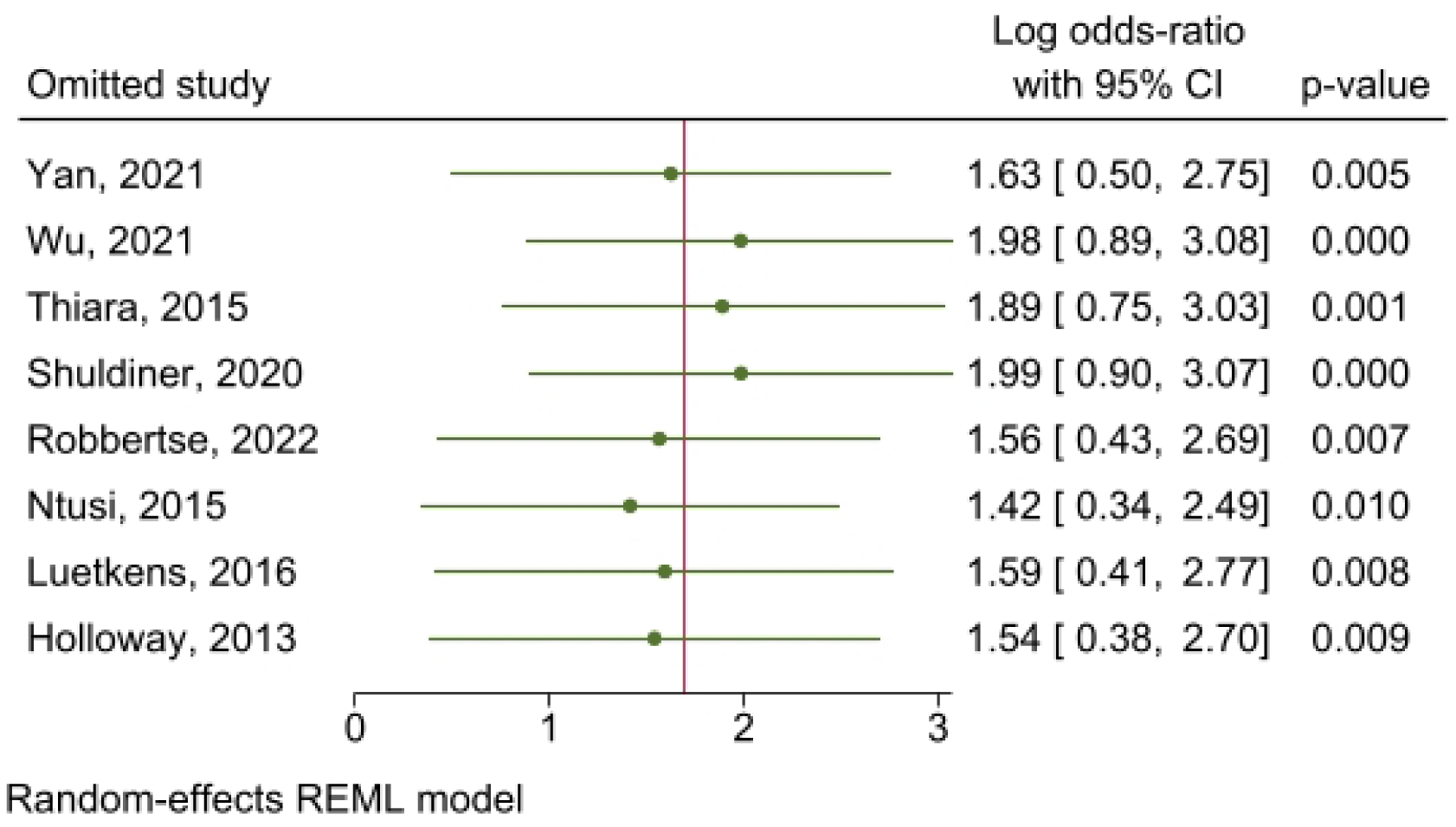
The results of leave one out analysis of LGE between PLWH and healthy controls.

**Supplementary Figure 2:**
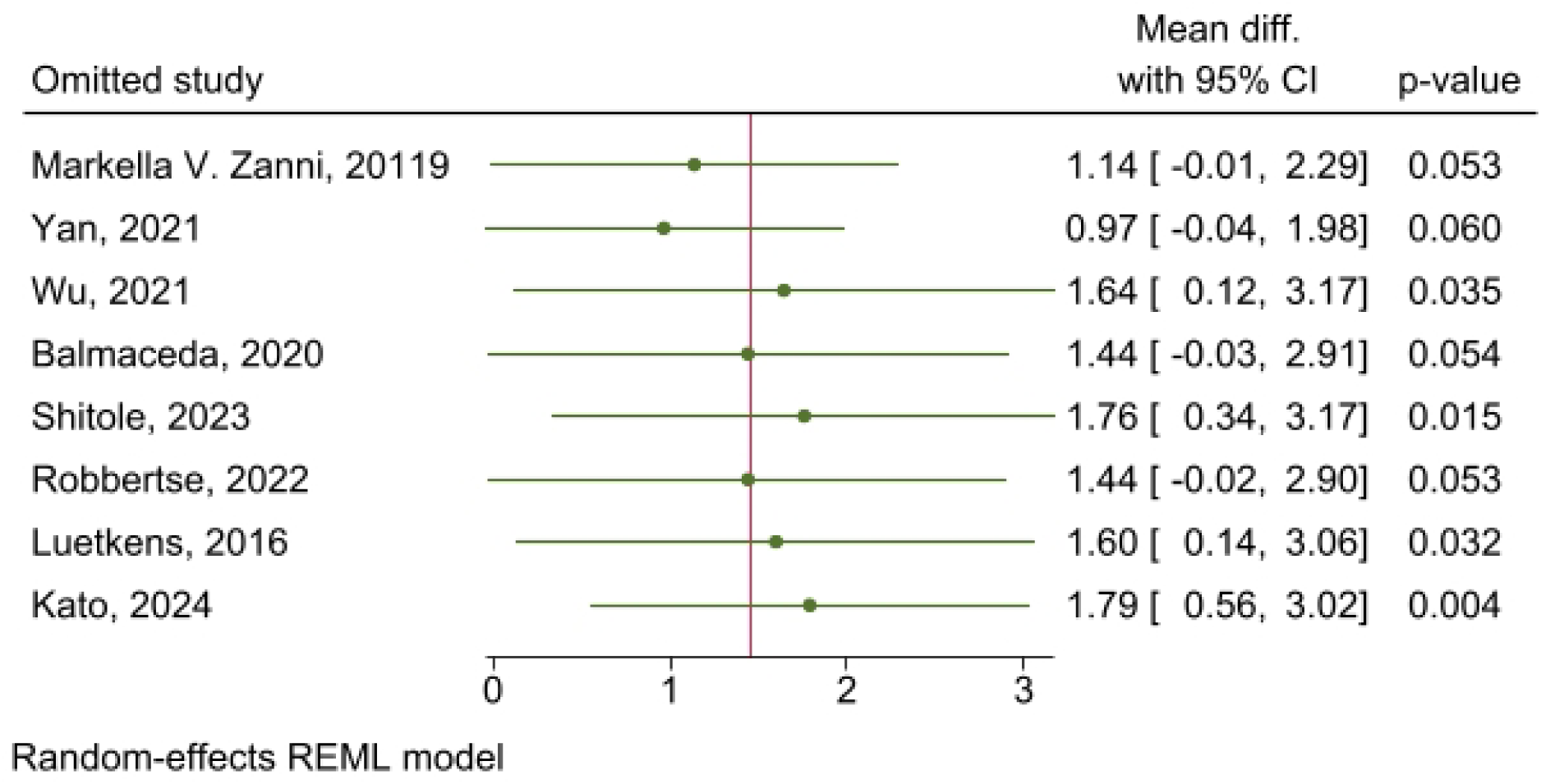
The results of leave one out analysis of ECV between PLWH and healthy controls.

**Supplementary Figure 3:**
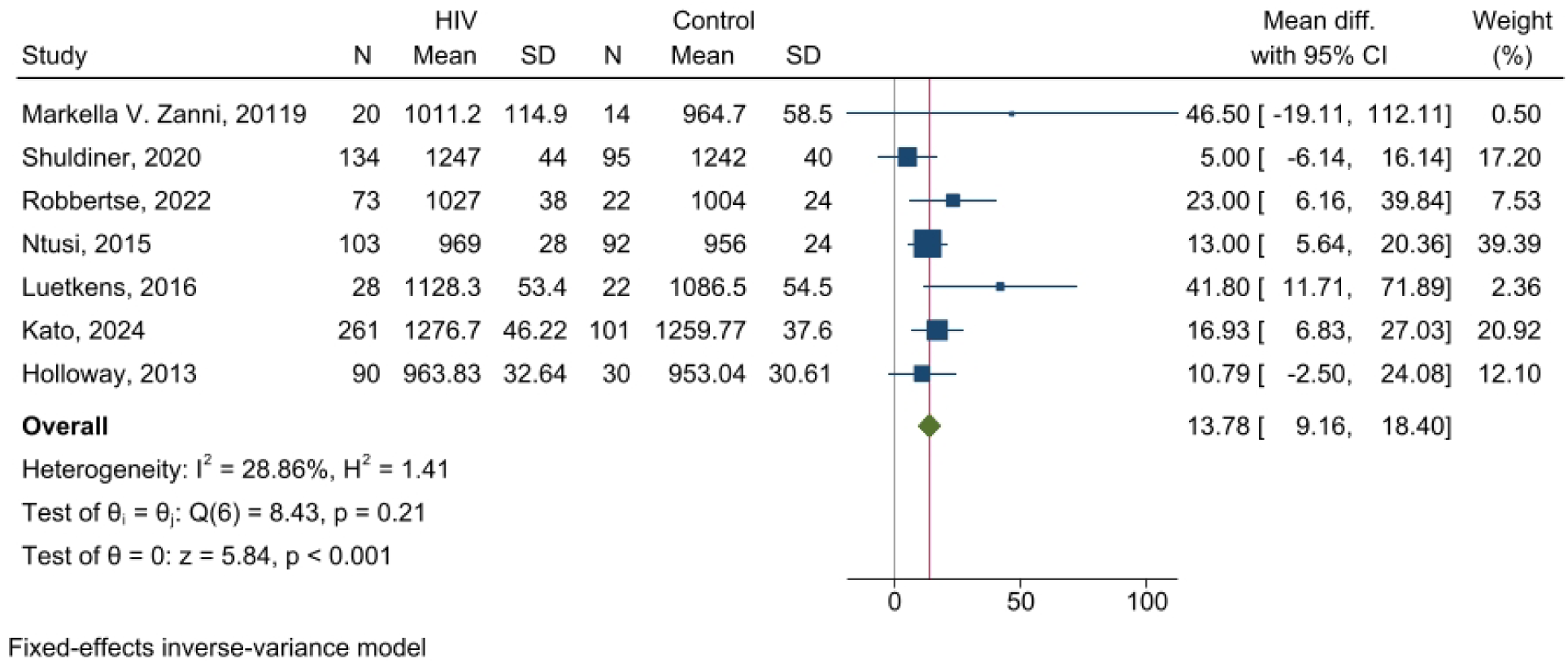
The results of pooled analysis of T1 between PLWH and healthy controls

**Supplementary Figure 4:**
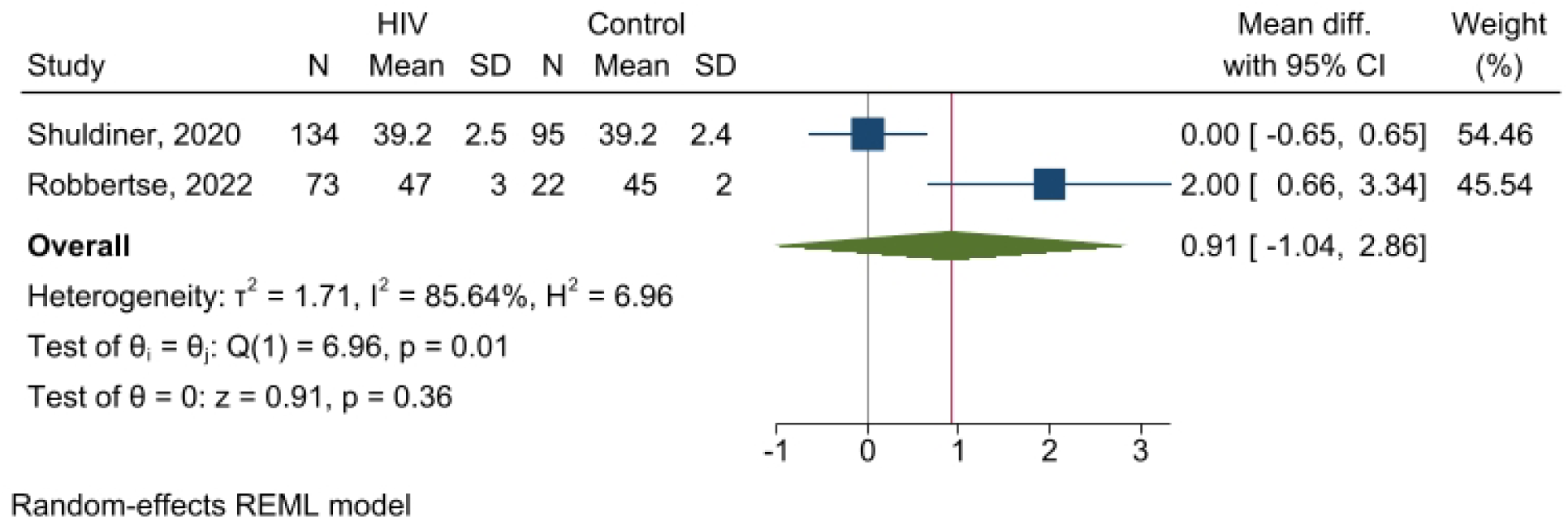
The results of pooled analysis of T2 between PLWH and healthy controls

**Supplementary Figure 5:**
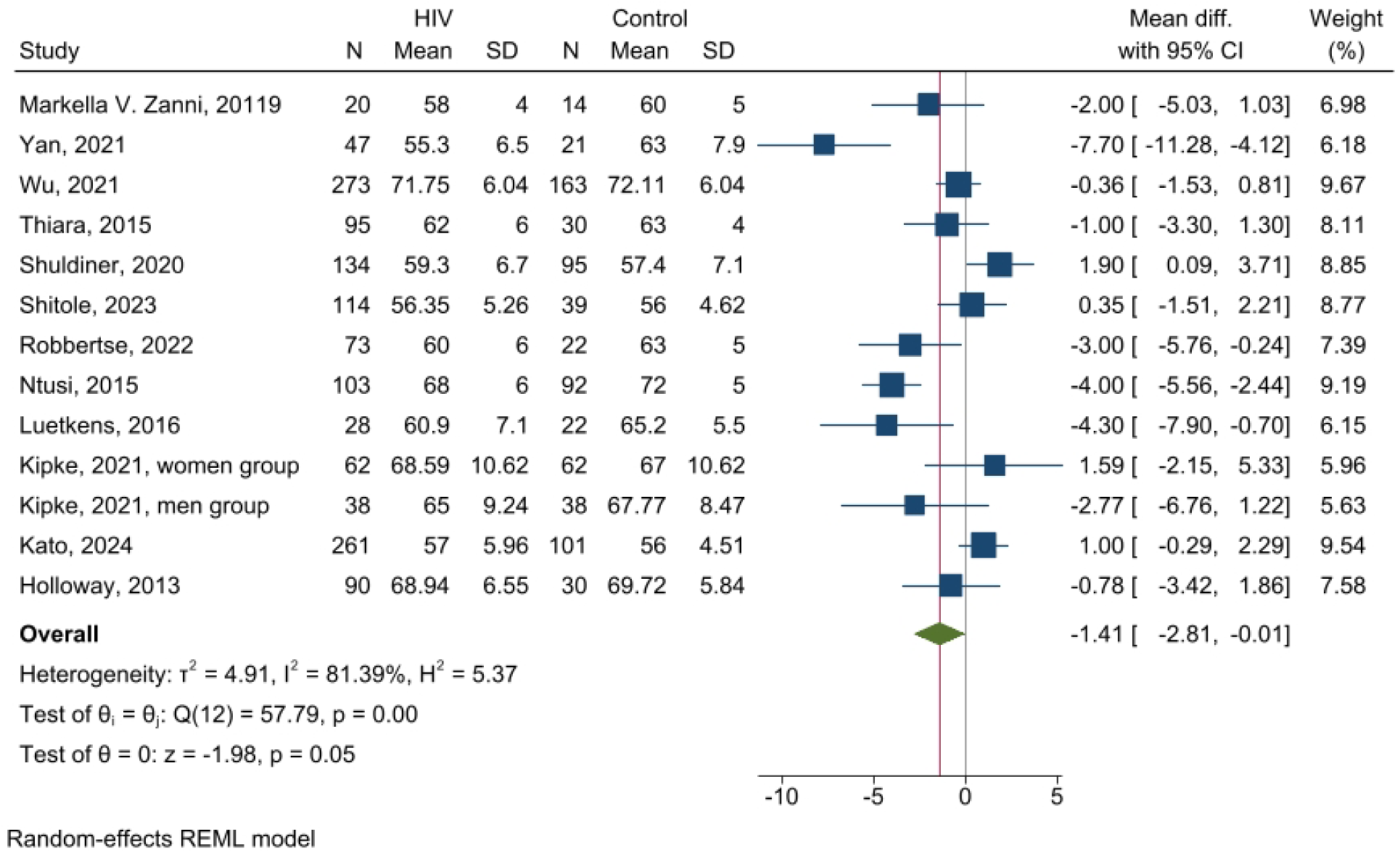
The results of pooled analysis of LVEF between PLWH and healthy controls

**Supplementary Figure 6:**
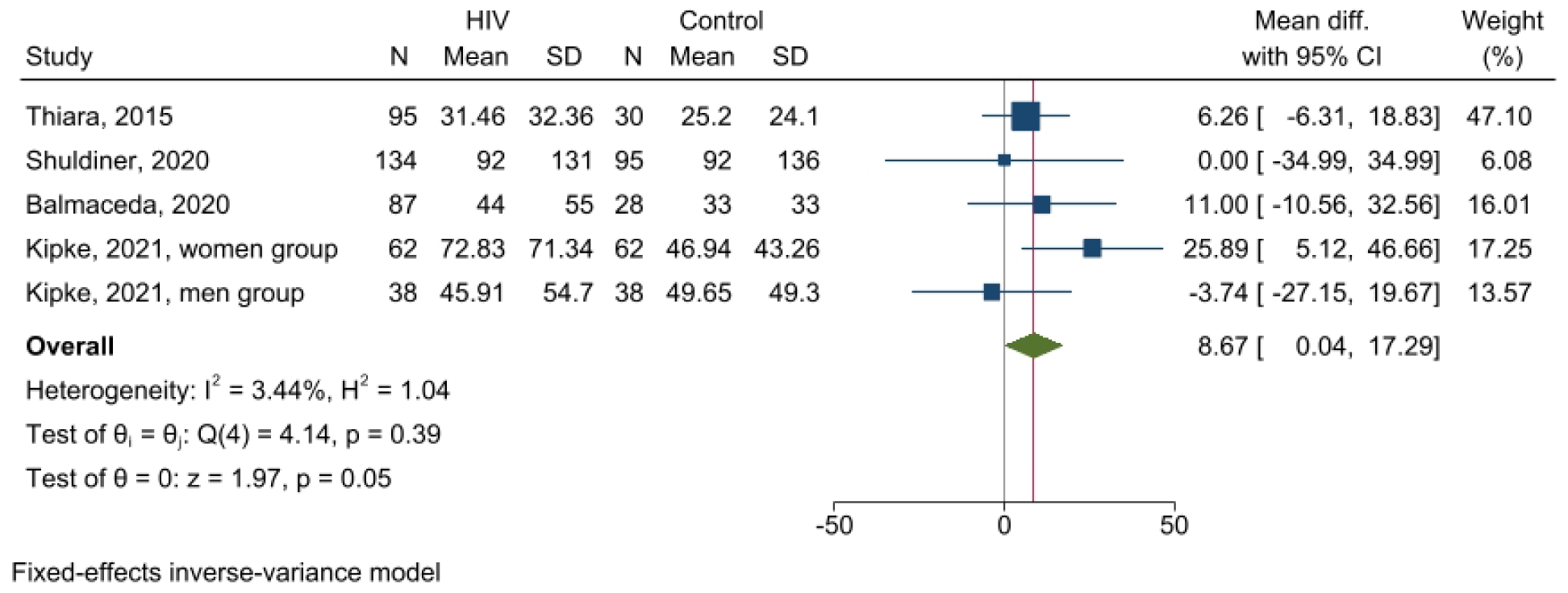
The results of pooled analysis of pro-BNP between PLWH and healthy controls

**Supplementary Figure 7:**
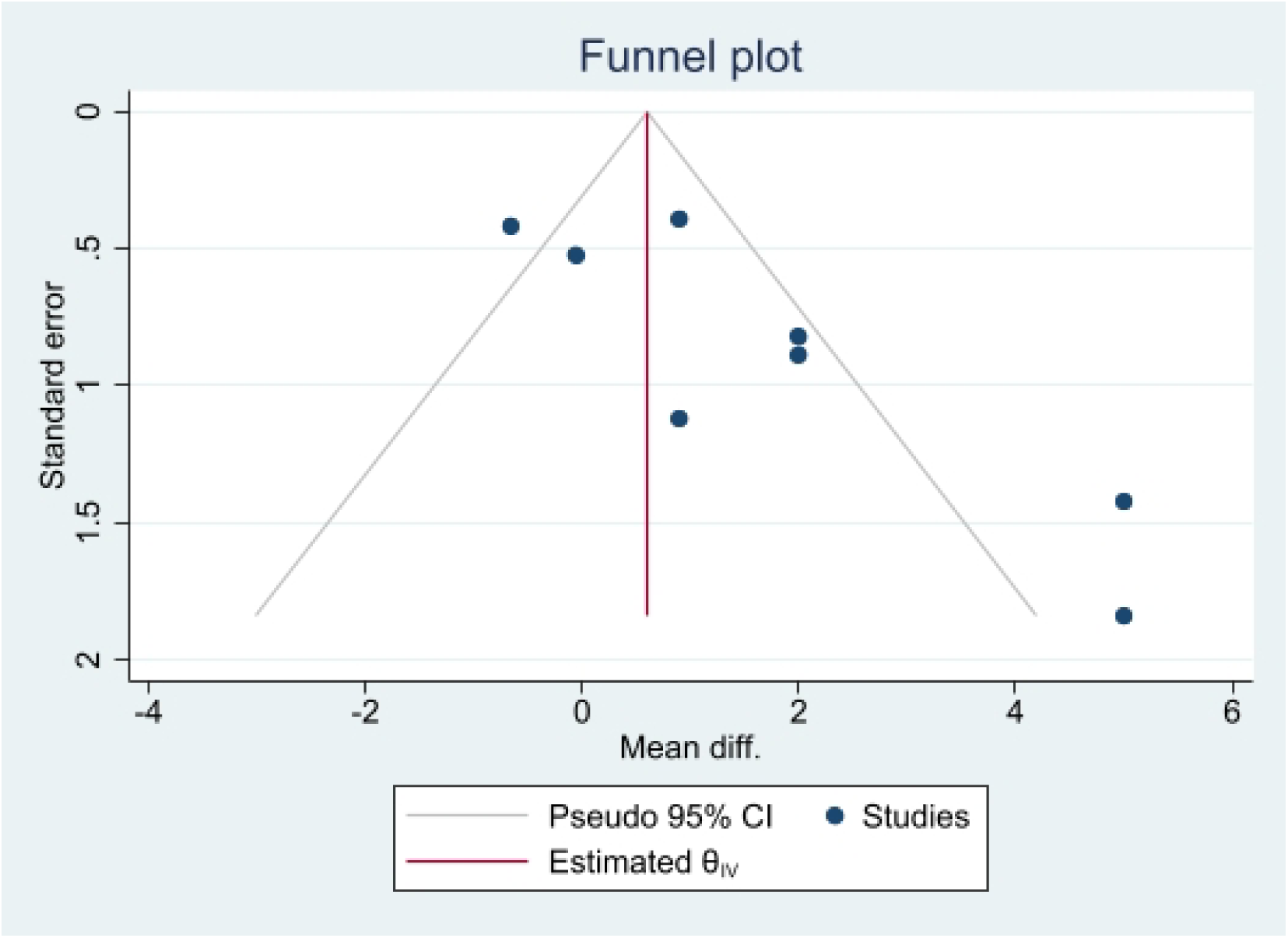
The funnel plot of studies reporting ECV in PLWH and healthy controls.

**Supplementary Figure 8:**
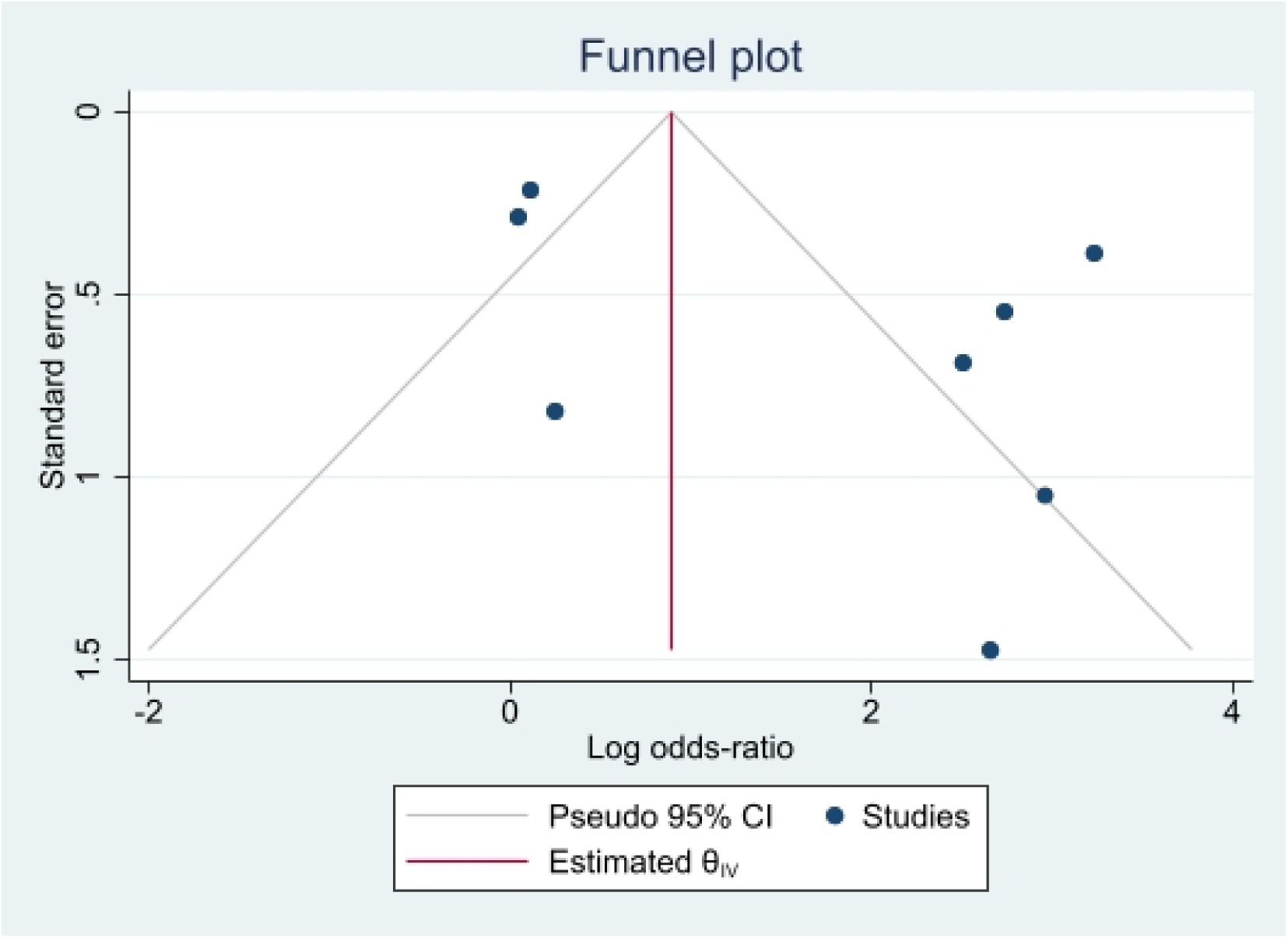
The funnel plot of studies reporting LGE in PLWH and healthy controls.

